# A new accessible adaptable COVID-19 model

**DOI:** 10.1101/2021.02.28.21252633

**Authors:** Michael G Baker, Maximilian de Courten, David A Sidloff, Alexander D L Baker, Walid S El Sayes, Faisal A Alaklobi, Abdulrahman S Alqahtani, Edward Fraser, Charles Cohen, Isam SM Osman

## Abstract

**Objectives:** Sophisticated epidemic models have been created to help governments and large healthcare organisations plan the necessary resources to manage the COVID-19 pandemic. Whilst helpful, current modelling systems are not widely accessible or easily adapted to different populations and circumstances. Our objective was to develop a widely applicable, easily accessible, adaptable model for projecting new COVID-19 infections and deaths that requires minimal expertise or resources to use. The model should be adaptable to different populations and able to accommodate social and pharmaceutical interventions as well as changes in the disease.

**Design:** A Susceptible, Infected and Removed (SIR) infectious disease model was created using widely available Microsoft Excel© software. The model is deterministic, generating projections based on the available data and assumptions made. It uses a process of *Monitored Forecasting* through *Visual Matching* of predicated vs observed curves to improve accuracy and facilitate adaptability. A review of the COVID-19 literature was performed in order to produce an initial set of adjustable parameters on which to base the output of the model.

**Setting:** This model can be adapted to different regions or countries for which the requisite input data (population size and number of deaths due to the disease) are available. This model has been successfully used with data from England, Sudan and Saudi Arabia. Data from NHS England were used for producing the illustrative results presented here. The model is a generic infectious disease forecast model which may be adapted to other epidemics.

**Intervention:** Governments, public health organisations, pharmaceutical companies and other public institutions may introduce interventions that affect disease transmission or severity. Other unknown factors such as new variants of the infective agent may do the same. The effects of changes in disease transmission are identified by the model when predicted and observed curves deviate. By aligning the curves an evaluation of the effect of the changes can be made.

**Outcome Measures:** The model graphically demonstrates projections for daily deaths, cumulative deaths, case mix (asymptomatic, symptomatic and severe infections requiring admission), hospital admissions and bed occupancy (ICU, general medical and total).

**Results:** The model successfully produced projections for the outcome measures using NHS England data. Users can adapt and continuously update the model correcting its projections as further local data becomes available. The Microsoft Excel platform allows the model to be used without expensive health information systems or computing infrastructure.

**Conclusion:** We present an SIR epidemic model that projects COVID-19 disease progression, is widely accessible, adaptable to different populations and environments as the disease progresses and is likely to be of benefit for identifying changing population healthcare needs.

## Introduction

First detected in December 2019, COVID-19 is now established worldwide with numbers of infections rising globally^1 2^. In April 2020 the World Health Organization assessed countries’ individual preparedness for the impact of COVID-19 and many countries were classified as “not operationally ready”^3^. Disease modelling may help prepare healthcare services by forecasting the likely number of cases and the associated resources required.

Mathematical models have been used to simulate scenarios and predict the spread and distribution of infectious diseases since the early 20th century^4^. Although the underlying concepts are often quite simple, mathematical formulations for these models can be complex especially when they aim to incorporate the potential impact of policies designed to limit the spread of the infection^5^. The availability of modelling expertise globally is limited, and many countries might not have the capacity to apply sophisticated models for their local health care planning needs. Furthermore, models that can be contemporaneously updated to assess the impact of changes such as non-pharmaceutical interventions (NPI’s) may be useful.

### Multiple models have been produced including

The Institute of Global Health^6^, (University of Geneva and the Swiss Data Science Centre, ETH Zürich-EPFL), COVID analytics^7^ the University of California (USA only)^8^, Los Alamos national laboratory^9^, The University of Washington^10^, the University of Texas (USA only)^11^ and Northeastern University (USA only)^12^.

Many are modified SIR models as is the model presented here. However, many rely on externally sourced data and assumptions not directly modifiable by the user for a local or regional setting. The data used in these models (frequently from China and Europe) can be from populations with different demographics, social contact patterns and healthcare systems and may therefore be poorly applicable to other parts of the world. Complex modelling systems with extensive data requirements also require modelling expertise, advanced computing technology, and may need established healthcare information systems that are not available in all circumstances where a model might be useful.

It was our objective to develop an accessible, easy to use model that can be periodically adjusted in order to improve tracking of current viral growth rates throughout the pandemic to aid healthcare planning including the need for hospital intensive care units (ICU) and general ward beds.

## Methods

We developed a modified Susceptible, Infected and Removed (SIR) model^13^. SIR models assume a fixed population^14^ that is divided into three groups, each representing a fraction of that population including; the Susceptible fraction (people yet to be exposed and infected), the Infected fraction (people who are contagious) and the Removed fraction (those who survived the infection and are assumed to be immune or those who did not survive). SIR models assume that the infection is spread directly from infected individuals to others; any non-immune individual after sufficient contact with an infectious individual will develop the infection and will be infectious to others (within a limited time period) and subsequent to this, that individual is immune^13^ (for a specified time period). SIR models are routinely used by epidemiologists to chart the evolution of epidemics over time. Within SIR based models, the infection initially spreads exponentially due to the low level of population immunity, and, as immunity increases the susceptible population decreases causing the rate of spread to decrease.

### Our model

Microsoft Excel© (Washington, USA) was chosen for the model platform as it is widely available and many people are familiar with its use. The model is deterministic in nature^15^. Data requirements are of two types (a) historic data on the population size and the dates of recorded deaths (and, if available, test and hospital admission data) and (b) assumptions about key characteristics of the disease. In the latter case the model is pre-populated with a set of generic parameters obtained from our literature search at the outset of the pandemic (Table 1). These can be used or replaced as local data becomes available. Using a simple visual mapping process, growth rates for the epidemic are derived and then used to project the further progress of the epidemic. As up-to-date data becomes available, the assumptions (particularly about transmission rate) can be further adjusted to produce updated projections.

**Table 1.** Model Assumptions. Table showing the initial generic assumptions and the initial assumptions for England.

*Model Assumptions*
| **Assumptions** | **Generic** | **England** |
| --- | --- | --- |
| Delay in symptoms appearing after infection, in days | 5 | 5 |
| Symptomatic cases as a percentage of all cases | 20% | 20% |
| Hospital admissions as a percentage of symptomatic cases | 20% | 12.5% |
| Hospital admission after symptoms in days | 5 | 12 |
| Admission length of stay in days | 14 | 7 |
| Percentage of general ward patients who do not survive | - | 31% |
| Admissions to ICU as a percentage of all admissions | 33% | 17% |
| Days after admission to hospital a bed is required on ICU | 14 | 7 |
| Percentage of ICU patients who do not survive | 50% | 53% |
| Average days after admission a patient dies | 14 | 7 |

Low testing capacity can be a limiting factor for many countries hence the number of positive tests may under-estimate the actual number of COVID-19 positive patients and adversely affect projections. We therefore selected deaths secondary to COVID-19 as a more accurate estimate of disease prevalence in order to better estimate epidemic growth rates.

Using the initial numbers of recorded deaths, the model first calculates the number of people infected at earlier time points that would be necessary to give rise to these recorded deaths. It then calculates each subsequent day’s infection by using the daily growth factor and the susceptible population adjustment factors indicated below to calculate the next day’s new infections and then repeats this process. As new death data becomes available, that data is used to re-calculate effective growth factors.

Growth factors derived from the initial levels of infection or death at the start of an epidemic are less reliable. Also, it is recognised that data collected may be influenced by factors other than disease transmission. Both of these influence the accuracy with which a growth factor can be determined. Our model therefore uses a weighted average of an early week’s inferred infection levels as the basis of subsequent calculations. As more data becomes available the graphical matching process is used to update and refine the calculated growth factors and projections.

### Growth rate calculations

This model utilises a calculated epidemic growth rate which is inherently linked to the basic reproductive number R_0_ which is more commonly described in the literature. The R_0_ may be defined as the average number of people a person will infect in total, during the entire time they are infective, in a population that is susceptible^16^. At the early stages of an epidemic a given level of R_0_ will result in an increasing number of new infections each day, with the number of new infections each day being a multiple of the number the day before (exponential growth). This multiple may be described as the Growth Factor^17^. This model determines an effective growth factor based on the assumption that each day’s infections are a function of the level of infections the day before.

The Growth Factor may be influenced by many variables including the infection itself, population characteristics, vaccines and non-pharmaceutical interventions (NPI’s). The growth factor calculated and used by this model incorporates the effect of such variables as data becomes available. The growth factor is applied to the susceptible fraction of the population which is assumed to reduce over time the more people become infected.

Subsequent new infections are projected using the adjusted growth factor applicable to each day as follows:

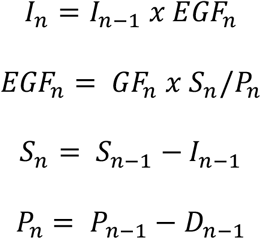

*I*_*n*_ *is the projected number of people newly infected on day n*,

*I*_*n*−1_ *is the number of people newly infected on day n-1 (the day before day n)*

*EFG*_*n*_ *is the Effective Growth Factor on day n, which is the factor by which the new infections are greater on day n than they were on day n-1*.

*It is the Growth Factor at that time (GF*_*n*_*) multiplied by the proportion of the population that is still susceptible to the infection*.

*GF*_*n*_ *is the Growth Factor on day n which can be expressed as the transmissibility of the infection, the average number of people an infected person will infect the next day if everyone they come into contact with is susceptible*.

*S*_*n*_ *is the number of the population that are susceptible on day n*

*P*_*n*_ *is the number/size of the population on day n*

*D*_*n*_ *is the number of deaths due to the infection on day n*

### Monitored Forecasting

Some environmental changes will be known (NPIs, vaccines etc), others will not (undetected new variants). Monitored forecasting maintains the continued utility of the model as time and circumstances change without the need to know the exact source of the underlying change. The model requires the user to input up-to-date data on deaths and if available tests and hospital admissions. By regularly monitoring the actual rates of infections and deaths against the model projections, through Visual Matching of the curves, model parameters can be improved whenever actual rates differ. By adjusting the model parameters in this way, the model’s projections remain useful and can demonstrate early changing rates of infection, whether the cause be known about or not.

### Visual Matching

In order to fulfil the requirement of a readily accessible and user-friendly model, we choose to incorporate a user-led, visual matching process which simplifies the adjustment of the model parameters. The model shows an overlay graph of the projected number of deaths compared to the observed number of deaths. The user then adjusts the growth factor or other assumptions so that the predicted deaths match those observed.

This process alerts the user to a change in growth rate (such as might happen with a new variant of the virus, or intervention), provides the user with an evaluation of an intervention’s effect and adjusts future projections, using the most recent growth rate.

### Modelling changing conditions

If the model’s graphical projections diverge from the graph representing the observed disease data, this suggests that either the value of an assumption(s) on which the model’s projection is calculated is inaccurate, or that its value has changed over time. If an initial assumption was wrong the value can be corrected to align the model projections with observed data. If the value of the assumption was initially correct but has changed as a result of the effect of new variables, such as vaccines or non-pharmaceutical interventions, a new value for the assumption can be changed from any specified date. In some cases, the date of the change will be known (e.g. introduction of social distancing measures) whilst in others it may not (e.g. the arrival of a new variant). Such scenario changes can be inserted at the time they occur or at appropriate dates in the model to better explain observed data.

The model also allows the user to insert hypothetical scenario changes to evaluate the potential impact of changed assumptions to test the possible effects of new NPIs, or new variants or other changes.

### Selecting parameters for the model

A review of the literature was performed in order to identify the generic parameters to be used in the model. All studies or national/international reports on COVID-19 were considered for review. Any duplicate data were excluded. PubMed databases (Medline, PubMed Central) were searched on 12 May 2020 for full text articles using the terms “COVID-19” OR “SARS-CoV-2” OR “Coronavirus” AND “outcomes” OR “mortality” OR “death”. The search was limited to clinical studies or case reports and only papers published in the last 6 months in order to exclude publications relating to other coronavirus. The reference lists of selected articles were also searched manually to identify relevant articles or reports. All authors independently reviewed the relevant articles to determine eligibility after which IO, MB and DS extracted data to obtain quantitative information for the following parameters: Delay in symptoms appearing after infection, Symptomatic cases as a percentage of all cases, Hospital Admissions as a percentage of symptomatic cases, Hospital Admission after symptoms, in days, Hospital Admissions as a percentage of all cases, Hospital Admission from date of diagnosis in days, Admissions to ICU as a percentage of all admissions, Average days a bed is required on ICU, Percentage of ward patients who do not survive, Percentage of intensive care unit patients who die, Mean days after admission a patient dies, Mean days after infection death occurs, Percentage of infected patients dying (including asymptomatic). Following this a virtual meeting was held during which authors agreed on a generic set of assumptions for the model (Table 1).

### Outcome

#### Model Projections

The model graphically demonstrates:

- Projected daily deaths.
- Projected cumulative deaths.
- Projected case mix - Case mix was defined as asymptomatic (no evidence of pneumonia), symptomatic (fever, cough, pneumonia) and severe infections requiring admission (including the need for oxygen, fluids or ventilation).
- Projected hospital admissions - admissions were based on the assumption that only severe infection would require admission to hospital.
- Projected bed occupancy - Bed occupancy was divided into ICU, general medical and total beds (ICU plus general medical beds).

## Results

The generic assumptions were updated for England as country-specific data became available (Table 1). In the UK this was largely influenced by the Semple and colleague’s publication using the ISARIC WHO Clinical Characterisation Protocol^18^. In England 12.5% of symptomatic patients required hospital treatment which was 2.5% of all COVID cases. We assumed that 17% of admissions to hospital would require ICU care and that 59%^19^ would not survive (1% of total infections) at 24 days after infections occurs.

We visually matched an initial daily growth factor to the England COVID-19 daily death rates reported by NHS England^19^. We visually matched the model’s growth factor to the number of daily deaths in England to estimate the actual growth factor (Figure 4).

**Figure 1.**
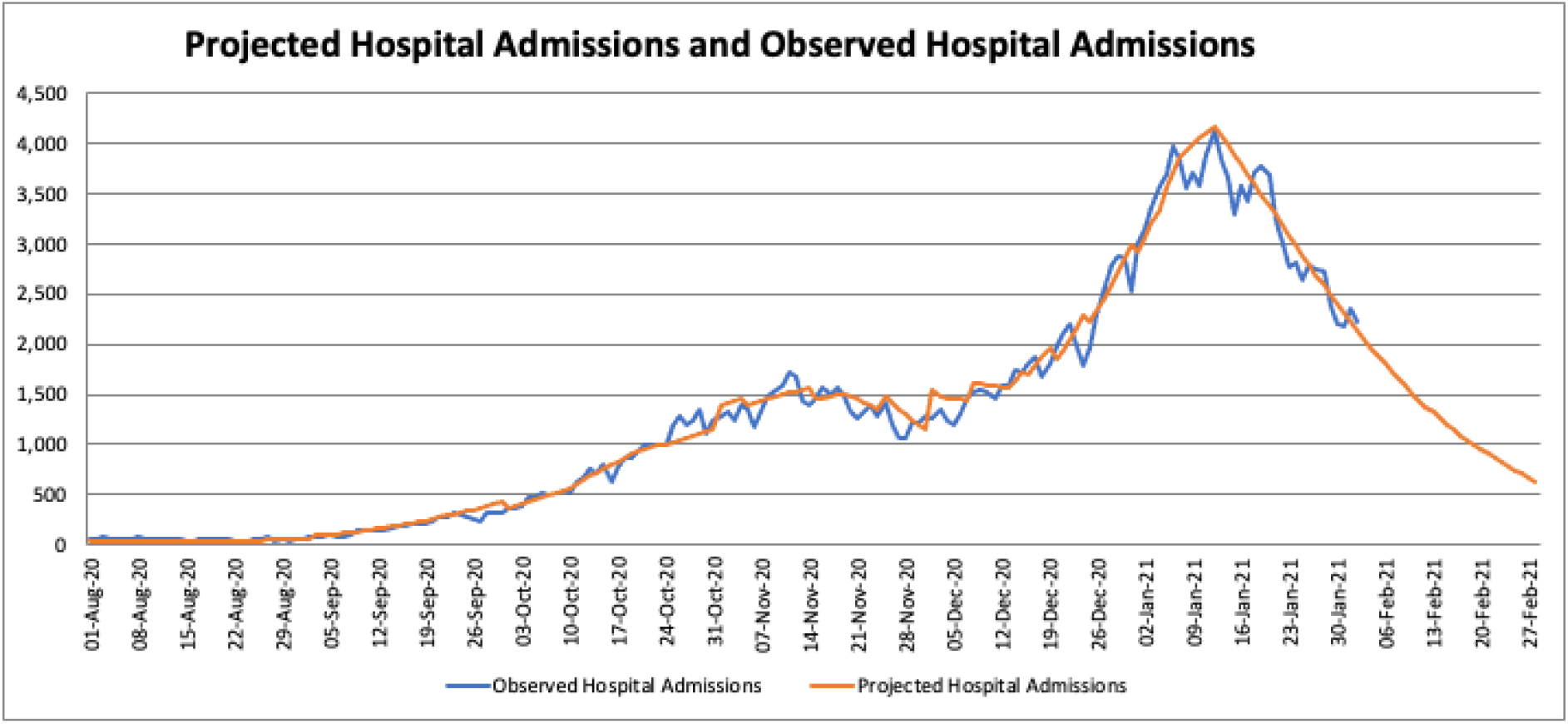
Figure showing overlay of projected hospital admissions and observed hospital admissions (NHS England Data) August 2020 – February 2021.

**Figure 2.**
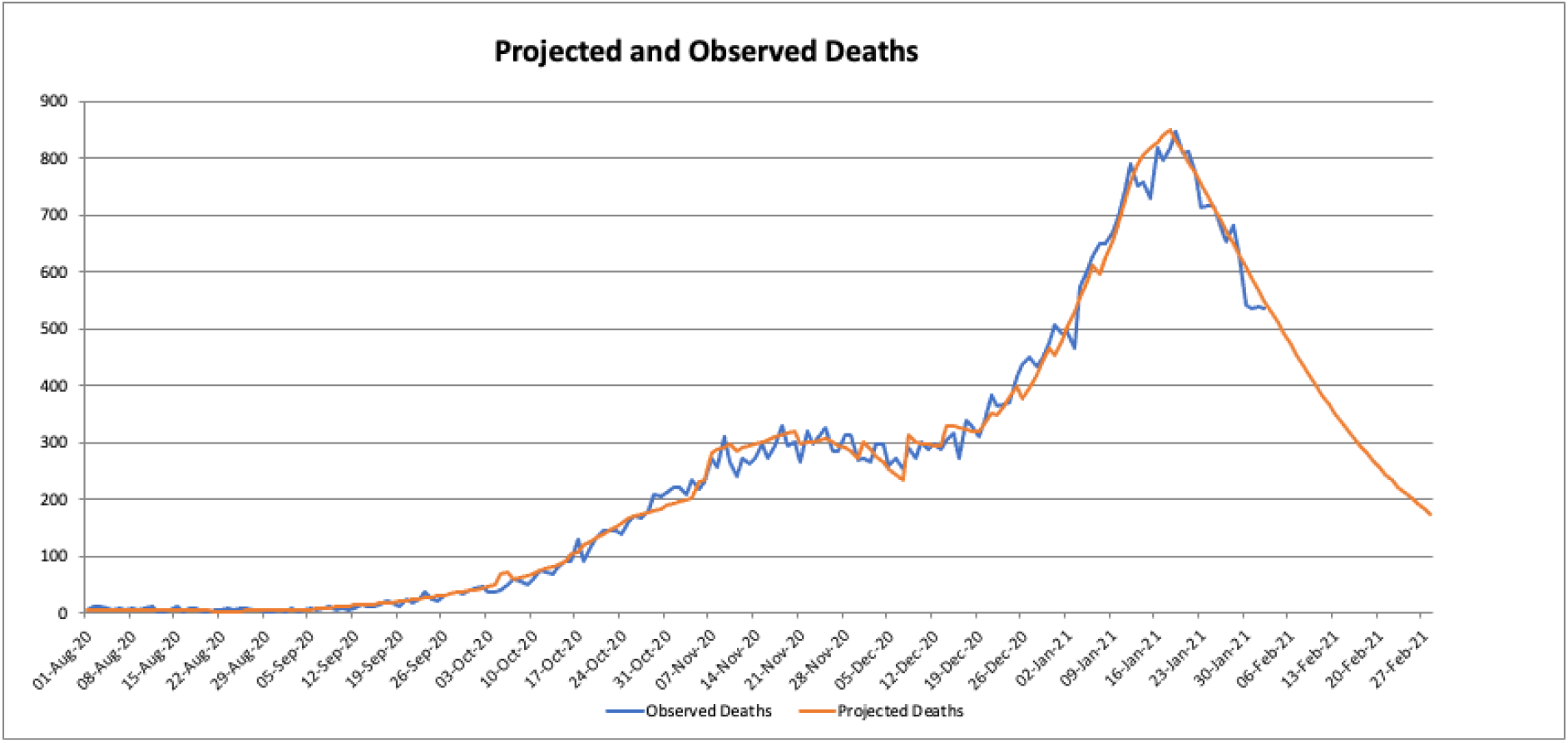
Figure showing overlay of projected deaths and observed deaths (NHS England Data) August 2020 to February 2021.

**Figure 3.**
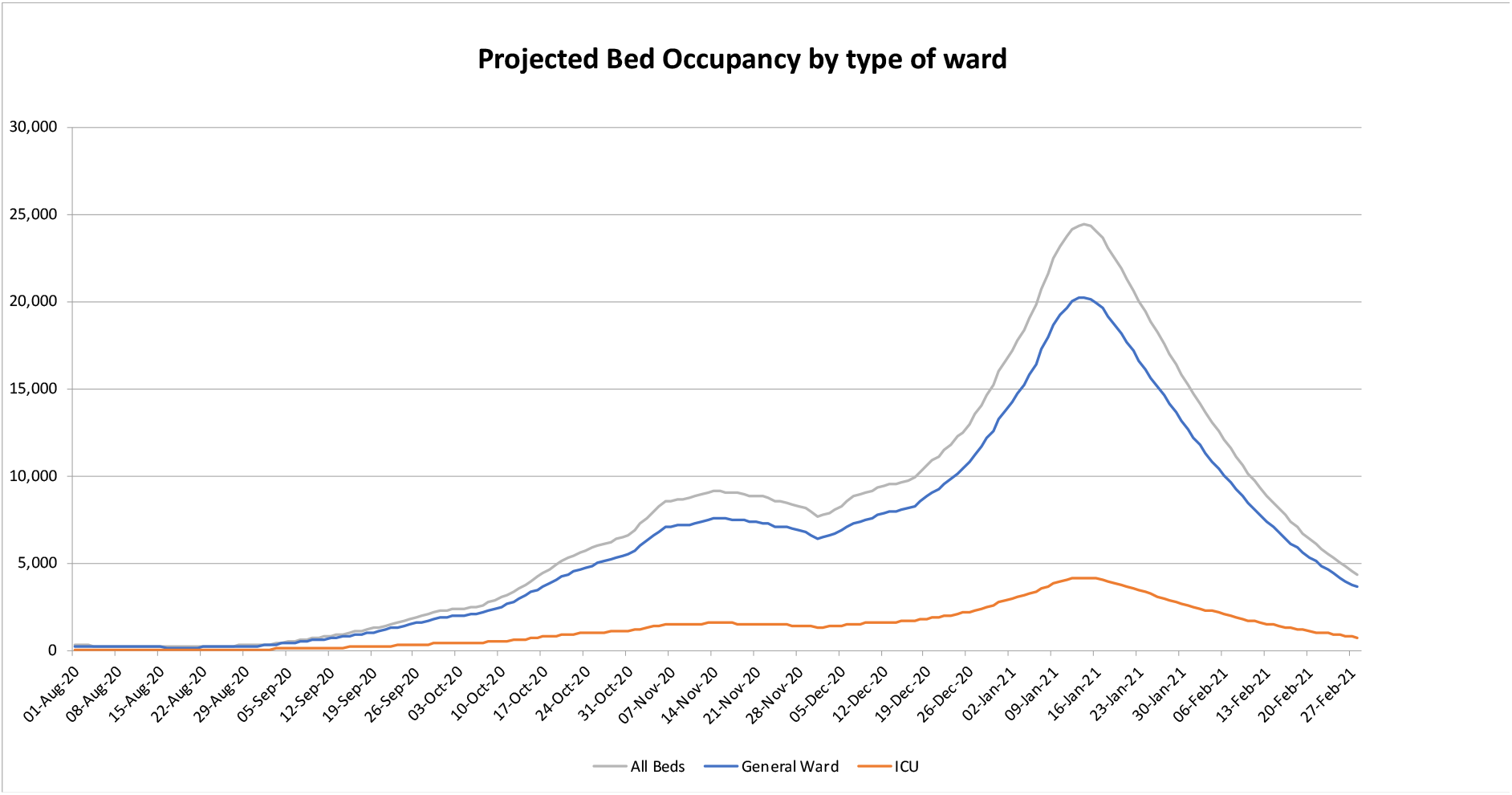
Figure showing projected bed occupancy by type of ward.

**Figure 4.**
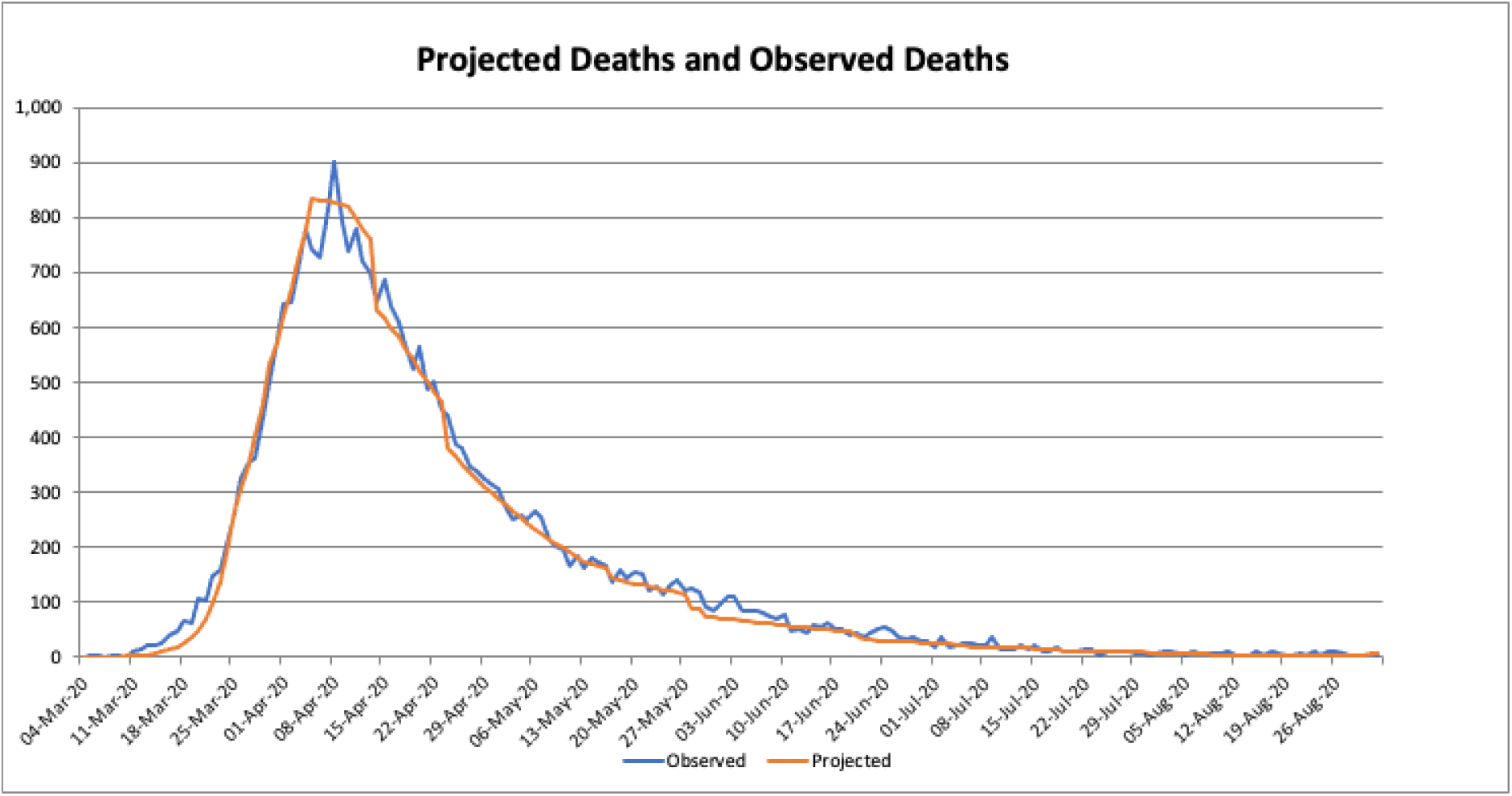
Figure showing projected and observed deaths for the period March to August 2020 (NHS England data).

Un-enforced (non-legally binding) social distancing guidance was issued on the 17^th^ of March 2020 in the UK (working from home, no non-essential travel), followed on 24^th^ of March 2020 with the introduction of legally enforceable social distancing guidance (monetary fines). The model initially projected approximately 1,900 patients per day requiring admission by the third week in March. Data to mid-April suggested that the growth factors had dropped indicating that the government’s interventions and publicity campaign were associated with a reduction in growth rate from 1.15 to 0.97 (Figure 5).

**Figure 5.**
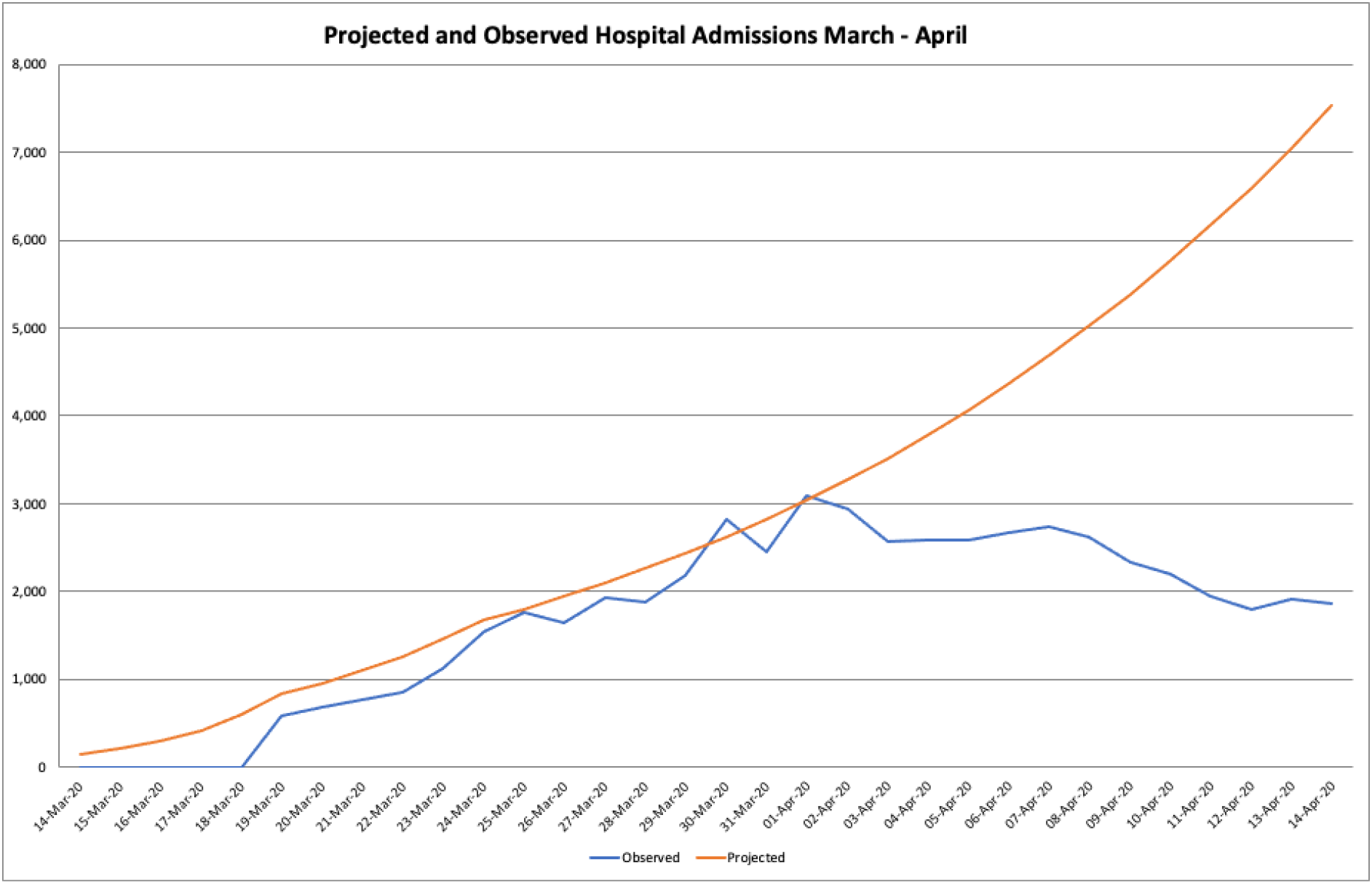
Figure showing diverging projected and observed hospital admissions (NHS England) as social distancing measures are introduced.

## Discussion

There are some limitations of an SIR based approach. One assumption of the SIR approach is that the population is fixed, however, populations are not entirely fixed even with the introduction of travel bans and movement restrictions. Another assumption is that after infection, a person is immune from the virus and therefore cannot be infected again (or pass the virus on). Although the ability to factor in temporary immunity exists in our model, we recognise this may not be the case and future research is required in order to fully understand the duration of immunity after infection with COVID-19 (or vaccination). None the less, SIR type models are a well-accepted methodology for projecting surge capacity requirements in viral epidemics. All models rely on the quality of data available. The data reported in this study utilises deaths in the hospital setting, however, England is a good example of how excluding data (e.g. out of hospital deaths or social care settings) can underestimate the true number of deaths. This is accentuated in the case of COVID-19 as it disproportionately affects the elderly who are more likely to die out of hospital.

Model projections are affected by the assumptions regarding the disease process on which the model is based. The generic assumptions applied to this model from our literature review are likely to be weak when applied to regions with divergent socioeconomics, demographics and healthcare access, as more often than not the published literature originates in high income settings. Use of the model requires basic proficiency in Microsoft Excel. Allowing the model assumptions to be adjusted introduces the possibility of user error. However, by allowing the adjustments the model can be adapted to better reflect different populations.

The British statistician George Box wrote “All models are wrong, but some are useful” ^20^. Models such as this cannot predict the future. The outcome of previously untested interventions or changing disease scenarios (such as the advent of new disease variants) cannot be known. Models such as this may be useful because as model data is updated the user is alerted to possible changes in scenario. They also facilitate the early evaluation of the effect of an intervention or change in disease scenario. Forecasts of local healthcare requirements can be made using model projections based on locally available data and assumptions. Different hypothetical disease scenarios can also be modelled by changing the model assumptions.

## Conclusion

An SIR model combined with *Monitored Forecasting* using *Visual Matching* has allowed the development of a novel, relatively simple to use COVID-19 model that allows the detection of early changes in growth rates and helps project the need for healthcare resource without the need for resource intensive health informatics infrastructure. This may allow a non-specialist to produce useful COVID-19 forecasts adaptable for their country, region or area of study.

## Data Availability

We confirm that we are happy share data related to this work.

http://epidemicprojections.org

## Supplementary information

This Microsoft Excel© based model can be downloaded from http://epidemicprojections.org/ at no cost.

No external funding was received for this work.

### Definition of terms

*In this paper we have used the terms Forecast and Prediction to mean an estimate of one possible outcome at a future time (based on judgements and the model projections)*.

*We have used the term Projection to mean a calculation of what might occur based on the data and assumptions used by the model*.

